# Prevalence, clinical profile and determinants of tungiasis among schoolchildren in Dschang Health District, Cameroon

**DOI:** 10.1101/2025.10.03.25337234

**Authors:** Earnest Njih Tabah, Djam Chefor Alain, Lélé Deffo Carole, Momo Anoumbo Ulrich, Guthe Kamdem Brice, Kenhale Zebaze Lunnelle Correcta, Nomedem Jaurès

## Abstract

**Background:** Tungiasis, a skin infestation caused by the flea *Tunga penetrans,* affects resource limited tropical communities of Latin America, the Caribbeans and sub-Saharan Africa. Recently considered by the WHO as a Neglected Tropical Diseases, epidemiological data to inform control strategy is lacking in Cameroon.

**Methods:** A cross-sectional study targeting schoolchildren from eight primary schools selected by stratified random sampling method was conducted from November 2024 to July 2025 in Dschang Health District, west Cameroon. Data was collected to assess the prevalence of tungiasis infestation, its associated factors, and related morbidity, while observing all necessary ethical requirements.

**Results:** Some 407 schoolchildren with a median age of 11 (7-16) years, participated in the study of whom 54.8% were males, 84.0% attended government primary schools, and 81.8% cohabited with domestic animals. Fifty-one of the schoolchildren were infested with tungiasis for an overall prevalence of 12.5% (95% CI: 9.5-15.9). Lesions were located mainly on toes (92.2%) hands (31.4%) and heels (27.%%), and morbidity included pains (72.5%), itching (66.7%), sleep disturbance (72.5%, lack of concentration (49.0%), deformation of toes (15.7%) and difficulties in walking (13.7%). Cohabitation with domestic animals (OR=5.15 (95%CI: 1.09-24.29), p=0.039) especially pigs(OR=2.34 (95%CI:1.13-4.82), p=0.022); being a pupil of class 5 (OR=22.11 (95% CI:4.73-103.36), p<001) or class 6 (OR=7.70 (95% CI: 1.97-30.11), p=0.003) were independently associated with tungiasis infestations.

**Conclusion:** This study reveals a substantial burden of tungiasis among schoolchildren in Dschang and associated with modifiable risk factors involving both the environment and individual behavioral factors. We recommend the implementation of integrated control strategies combining animal reservoir control, hygiene education, and adoption of WHO recommended tungiasis treatment practices. More research is needed to understand the wider spectrum of tungiasis in Cameroon.

**Author summary:** Tungiasis, commonly known as jigger, is caused by a parasitic flea and mostly affects people living in poor communities of Latin America, the Caribbeans and sub-Saharan Africa. As jiggers have recently been considered a public health problem by the WHO, there is not enough information on the disease in Cameroon.

We therefore carried out a study to assess the burden of the diseases among primary schoolchildren from selected government and private confessional schools in Dschang health district as part of efforts to understand its situation in Cameroon.

Among the 407 schoolchildren surveyed, 51(12.5%) of them were infested with jiggers. Toes, hands and heels were the affected parts of the body, and led to pain, itching, sleep disturbance, lack of concentration, deformation of toes and difficulties in walking among the infested schoolchildren. We found out that the schoolchildren took jiggers for granted and practiced self-extraction of lesions followed by application of some ointments and kerosene as means of treatment. We discovered that jigger infestation was closely associated with cohabitation with animals, especially pigs, and being a pupil of class 5 or 6.

We conclude that jiggers constitute a major problem among schoolchildren in Dschang and is associated with modifiable risk factors. We therefore recommend the limitation of direct cohabitation with animals, hygiene education and the adoption of the WHO recommendations for the treatment of jiggers. We also recommend more research to understand the wider spectrum of tungiasis in Cameroon.

## Introduction

Tungiasis, a painful skin infestation caused by the flea *Tunga penetrans*, is a major health challenge in remote and poor tropical communities of the South America, the Caribbean and Sub-Saharan Africa(1–6). It is considered a disease of the poor(7) and has recently been classified as a neglected tropical disease (NTD) by the WHO(8,9). This neglected tropical disease disproportionately affects children and the elderly(10–12). In sub-Saharan Africa, a pooled prevalence of 33.4% was reported in the general population in 2020(1), while a pooled prevalence of 37.86% was reported among school-aged children in 2025 (2). Alarming high prevalences have been reported among school-aged children in the northwest of Cameroon (60.5% - 72.1%) (11), Ethiopia (58.7%) (13), Southern Nigeria(14) and Kilifi – Kenya (48%)(15). The complex transmission cycle of this parasitic disease involves contaminated soil where eggs hatch and develop into adult fleas thrive (7,16) and either the humans, domestic or wild animals where the adult female flea embeds itself for the purpose of developing eggs, that are expelled into the soil to complete the cycle(17,18). Many risk factors for the transmission of tungiasis have been identified including: living in houses with earth floors, cohabiting with domestic animals, not wearing shoes at all or wearing them irregularly, and poor personal hygiene (1–3,13,19). Clinical manifestations of the tungiasis occurs within few hours from the penetration of the female sand flea and increases in intensity and severity as the flea grows. They include itching, swelling, pains, and ulceration(2,5,7,10,18). Severe complications, frequently observed in endemic areas, range from bacterial superinfections, through tetanus, to permanent physical deformities of the toes and feet particularly affecting patient mobility(1,2,7,17,18,20). Furthermore, sleep disturbance, lack of concentration, school absenteeism and poor performance at school have been found to be associated with tungiasis among schoolchildren(21,22). In Cameroon, there is paucity epidemiological data, with less than five studies on tungiasis that have revealed worrying prevalence rates of 32.7% in Bangou in the West region(23) and 53% in the Northwest region of the country(11). The Dschang health district, an agro-pastoral rural zone in West Cameroon, presents a potential risk for tungiasis transmission, however, no study has been conducted there to date to establish its veracity. Our research aims precisely to fill this gap by pursuing three major objectives: to determine the current prevalence of tungiasis among schoolchildren, to describe the clinical profile, and to identify the main socio-environmental factors associated with its transmission in Dschang health district.

## Methods

### Study setting and design

We conducted a cross-sectional study among primary schoolchildren in selected schools in Dschang health district (DHD), one of the 20 health districts of the West Region of Cameroon (Fig 1) from April to May 2025. Dschang is situated 339km northwest of Yaounde, the capital of Cameroon. The district is subdivided into 22 health areas of which five are urban and 17 rural, all covering 322 villages. There are 383 primary schools (286 government schools and 97 private schools) in the district. The health district has a population of 248 528 inhabitants (126749 females and 121779 males). Majority practice subsistence farming and rearing animals (pigs, goats, fowls, and domestic pets like dogs and cats).

**Fig 1.**
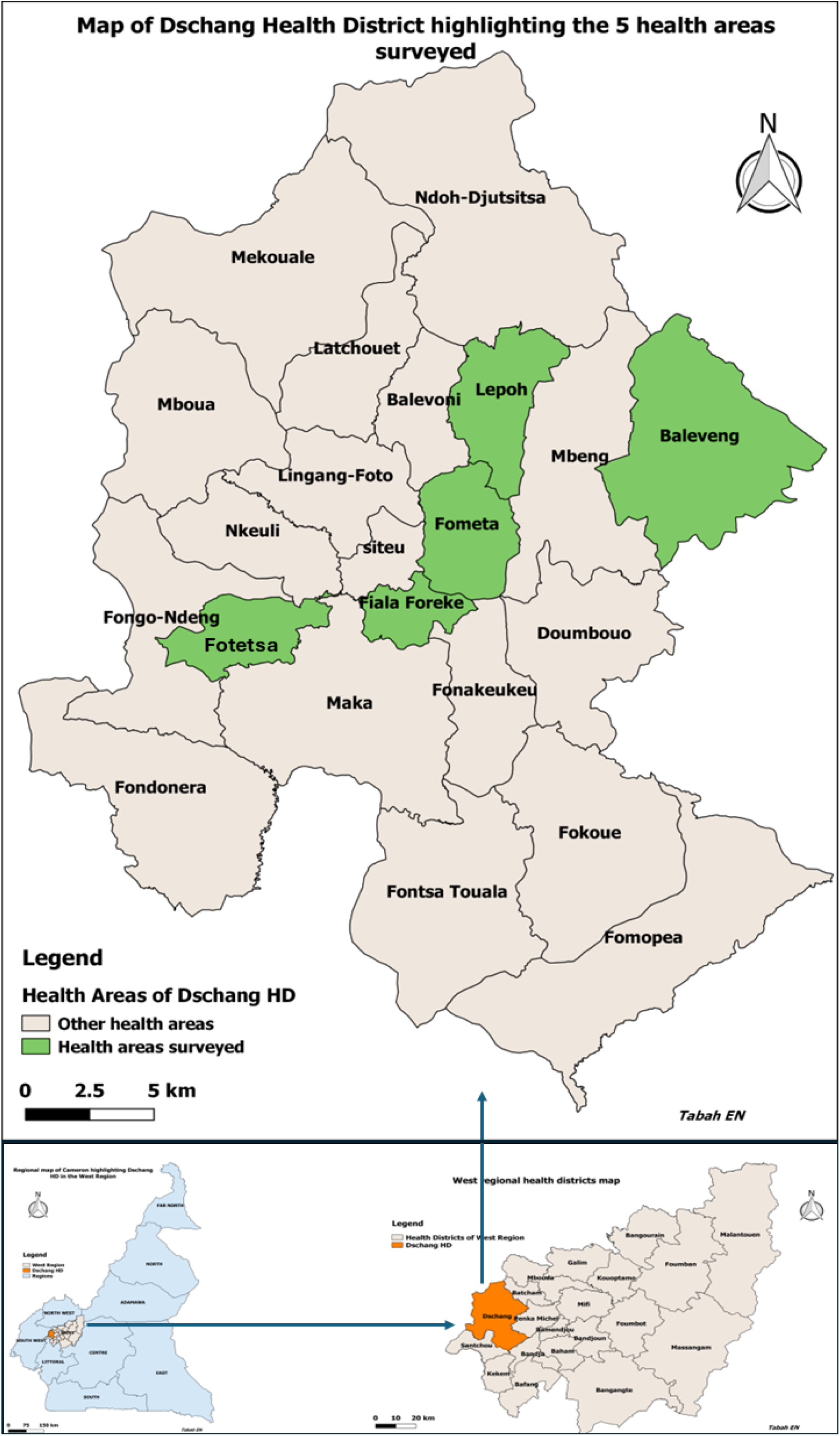
The map of Dschang Health District: highlighting the 5 health areas in which the study was carried out. The Dschang health district is one of the 20 health districts of the West Region of Cameroon. It is subdivided into 22 health areas of which five are urban and 17 rural, all covering 322 villages, and 317 schools.

### Study population and sampling procedure

The study targeted primary schoolchildren aged 7 to 15 years in Dschang health district. The sample size calculation, based on the Cochran formula, considered an anticipated prevalence of 32.7% reported in Bangou, west Cameroon [4], a margin of error of 5% and a non-response rate of 15%, resulting to a minimum sample size of 389 participants.

The sampling strategy combined a stratified multi-stage cluster approach. First, we stratified the district into urban and rural health areas, then randomly selected five health areas (two urban and three rural). In each selected health area and based on population weight, one or two primary schools were randomly selected. Finally, eight primary schools were selected including six government schools and two private confessional schools (Table 1). All the school buildings were divided into several classrooms. The classroom floors of the six government schools and one private confessional school had cracked concrete floors meanwhile the classroom floors of one private confessional school had smooth concrete floors (Fig 2). The total sample size was then proportionately allocated to each of the selected schools. Participants were chosen among schoolchildren of classes 3-6 present on the day of the survey in each selected school through systematic random sampling.

**Fig 2.**
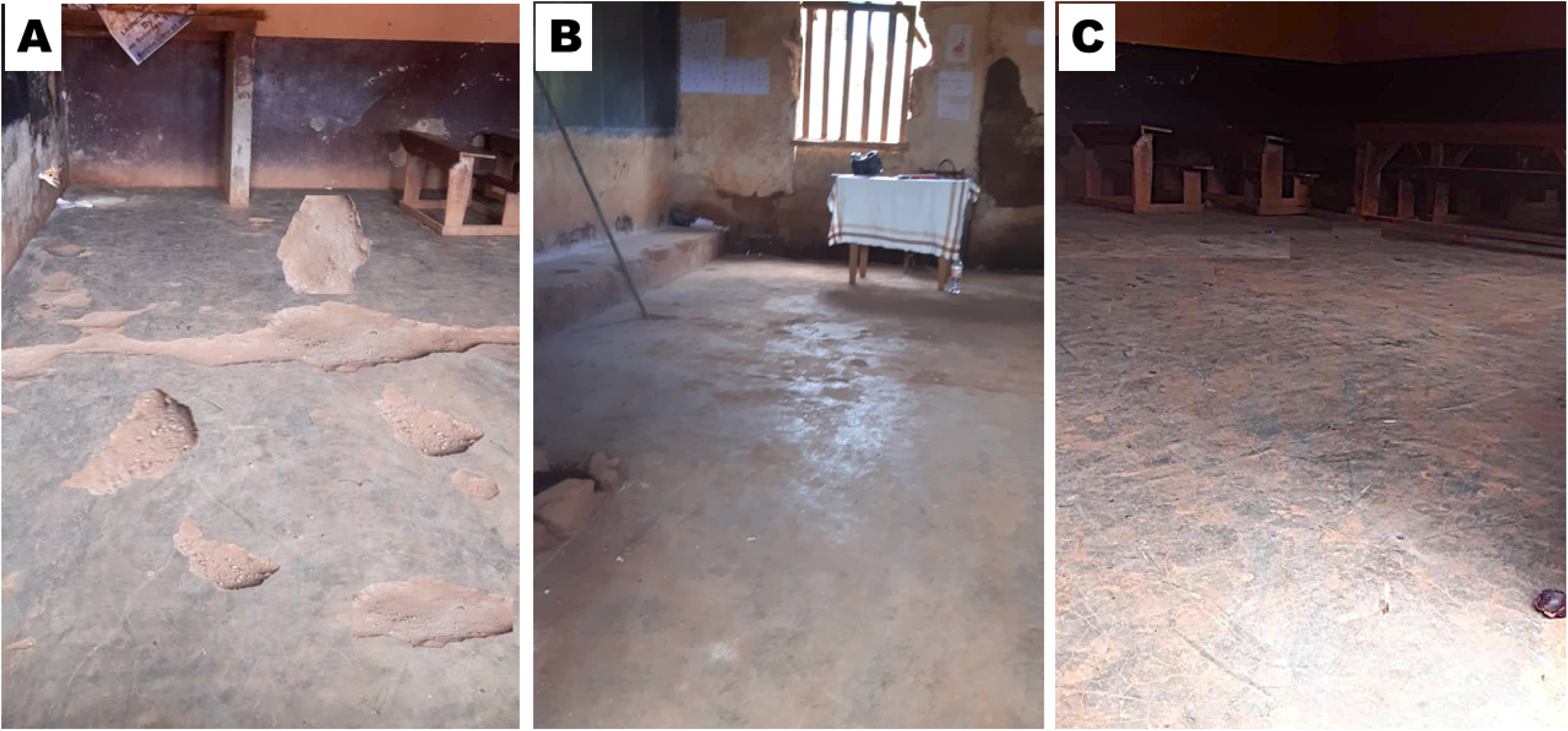
Classroom floors of selected schools: A: Cracked concrete (Baleveng central government school), B: moderately cracked concrete (Fiala-Foreke urban government school), C: smooth concrete (St Albert Catholic school Fiala-Foreke). Pictures by Lele Deffo Carole.

**Table 1.**
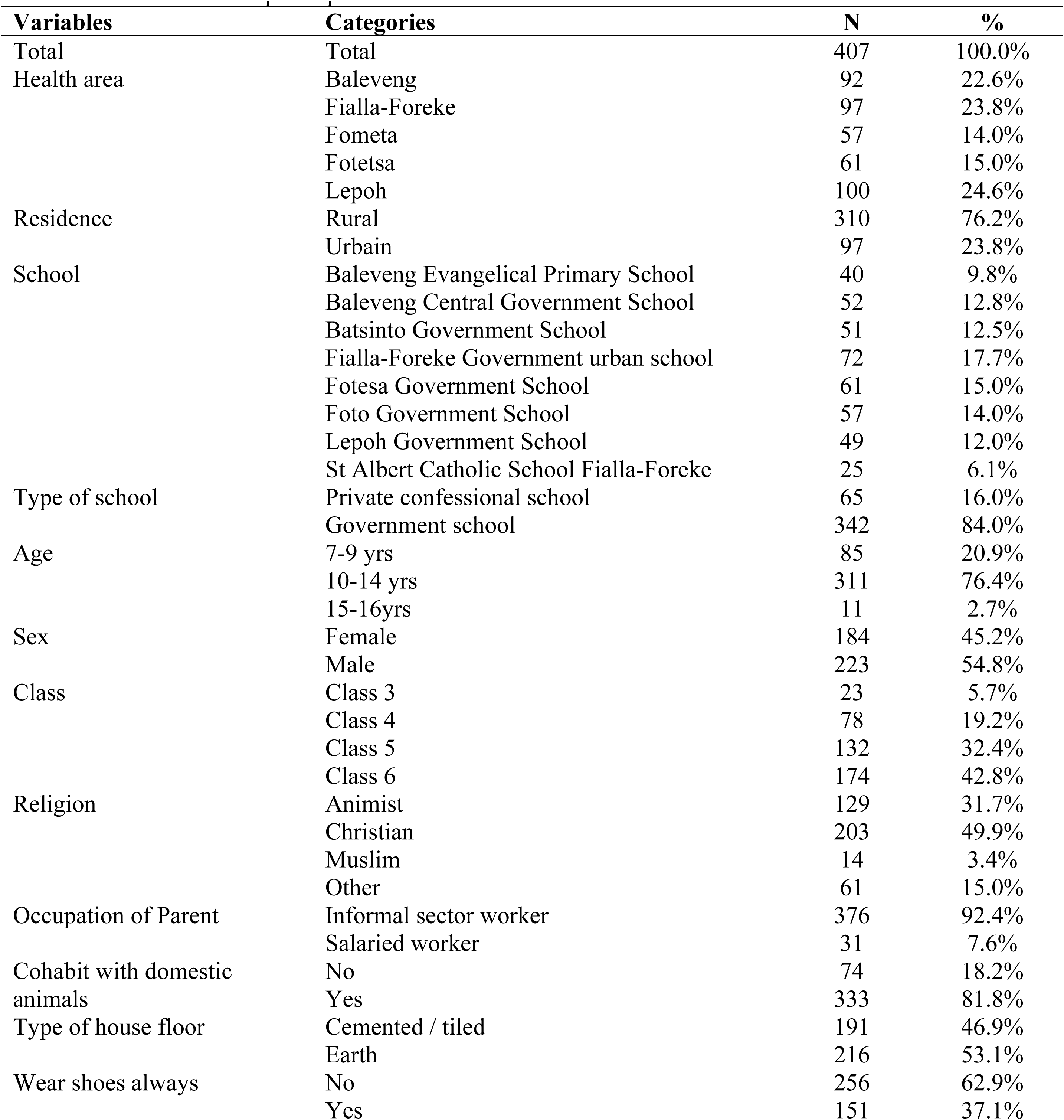
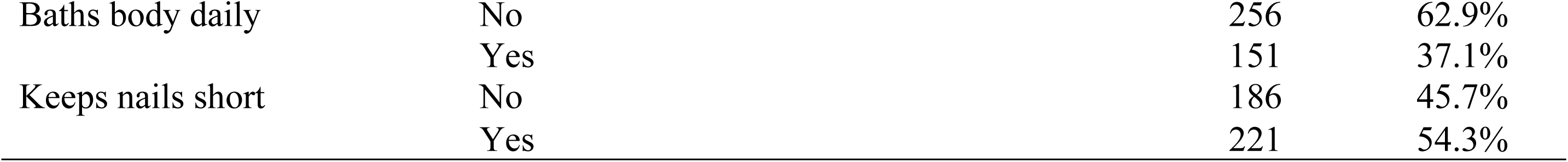
Characteristic of participants.

### Data collection and handling

Data collection included two main components: a standardized clinical examination and the administration of a structured questionnaire. A trained team of two students (one medical student and one master’s student in public health) collected data on the field. Prior to data collection, the team visited each school two days before to present the ethical clearance and administrative authorization from the divisional delegation of basic education to the school authorities and sensitize them and the pupils on the survey. Then after, they distributed flyers on the survey and consent forms to the pupils to take to their parents to sign and return to them in preparation for the survey. When the team came back two days after, only pupils whose parents had given consent and who were also willing to participate in the study were recruited.

Clinical examination was performed on the schoolchildren to evaluate infestation with *Tunga penetrans* according to the procedure described by Feldmeier et al. (2004)(24). Then after, a pre-tested questionnaire was administered face-to-face and collected information on sociodemographic characteristics, living conditions (type of house floor, presence of domestic animals), hygiene practices (frequency of bathing, nail care), risk behaviors (walking barefoot, self-extraction of lesions).

The data were anonymized and stored in secured place until they were analyzed. Only the members of the survey team had access to the data.

### Study variables

Infestation with *Tunga penetrans* was considered as the dependent variable. Sociodemographic characteristics (Age, sex, health area, residence, school, class, occupation of parents, religion); living conditions (type of house floor, presence of domestic animals: pig, fowls, goats, dogs and cat); hygiene practices (shoe wearing of habit, nail care habit, and frequency of bathing); risk behaviors (walking barefoot, self-extraction of lesions) were used as independent variables.

### Operational definitions

**Infestation with *Tunga penetrans***: based on the Fortaleza Classification, any schoolchild who presented with any of the following: a viable flea stick to the skin; a dark itchy spot in the epidermis; a white nodule of 3-10mm of diameter with a central black dot; a brownish-black circular crust with or without swelling and/or pus; circular residue in the keratin layer of the sole of the foot or irregular thickening of the nail rim or loss of toenail (24), upon clinical examination was considered pathognomonic. All such lesions were counted and recorded for each child examined.

**Intensity of tungiasis infestation:** the intensity was classified as mild (1-5 lesions), moderate (6-30 lesions) and severe (>30 lesions) according to the classification suggested by Muehlen et al(25).

### Statistical analysis

Data was entered into Microsoft Excel (Microsoft Office Professional Plus 2021) and then exported to SPSS software version 20.0. (IBM SPSS Statistics 20.0, IMB Corporation, 2023) for analysis. Means and percentages were analyzed for continuous and categorical variables. Pearson chi-squared and Ficher’s exact tests to verify the existence of associations between variables in univariate analyses, with level of significance set at p-value < 0.05, for 95% confidence interval. Then binary logistic regression using a forward stepwise likelihood inputting presence of tungiasis or not as dependent variable and the socio-demographic, environmental and behavioral factors whose levels of significance were less than or equal to 0.25 in the bivariate analyses as independent variables. Odd ratios (OR) and their 95% confidence intervals were calculated, with significance threshold set at p < 0.05.

### Ethical considerations

The study received approval from the West Regional Ethics Committee for Human Health Research (N° 427/30/04/2025/CE/CRERSH-OU/VP), and administrative authorization from the West Regional Delegation of Basic Education (N° 179/25/L/MINEDUB-O/SDAG/IMS). Informed consent for the study was obtained from parents of participating schoolchildren and school principals of participating schools. Then assent of the schoolchildren participating in the study was sought. Data confidentiality was strictly preserved by anonymizing the questionnaires.

## Results

A total of 407 primary school children participated in the study, recruited from 8 primary schools located in 5 health areas. Details of their characteristics are given in Table 1.

About 76% of the children were recruited from schools situated in rural health areas while close to 24% came from schools in urban health areas. Eighty-four percent (84%) were from government schools and 16% from private confessional schools. The age range was 7-16 years, Majority of the schoolchildren (76.4%) were aged 10-14 years, and 54.8% were males. Children from class 6 constituted majority of participants with 42.8% followed by class 5 and class 4 with 32.4% and 19.2% respectively. Close to 82% of schoolchildren lived in houses where they cohabited with domestic animals, and where 53.1% of the floors were earthed. About 63% of the children did not wear shoes always, and the same proportion did not take daily baths. Only 45.7% of them kept their nails tidy.

### Prevalence of tungiasis among primary schoolchildren

The overall prevalence of tungiasis among the schoolchildren surveyed was 12.5% (95% CI: 9.5% - 15.9%). Table 2 shows detailed distribution of the prevalence of tungiasis by socio-demographic and environmental variables.

**Table 2:** Distribution of tungiasis in relation to socio-demographic, environmental and behavioral factors.

| Variables | Categories | N | Prevalence of tungiasis |  | 95% CI of prevalence |  | P-value |
| --- | --- | --- | --- | --- | --- | --- | --- |
|  |  |  | n | % | Lower | Upper |  |
| **Overall** | **Overall** | **407** | **51** | **12.5** | **9.5** | **15.9** |  |
| Health Area | Baleveng | 92 | 22 | 23.9 | 15.6 | 33.1 | **<0.001** |
|  | Fiala-Foreke | 97 | 17 | 17.5 | 10.6 | 25.7 |  |
|  | Fometa | 57 | 1 | 1.8 | 0.04 | 6.4 |  |
|  | Fotetsa | 61 | 4 | 6.6 | 1.8 | 13.9 |  |
|  | Lepoh | 100 | 7 | 7.0 | 2.9 | 12.7 |  |
| Residence | Rural | 310 | 36 | 11.6 | 8.5 | 15.8 | 0.317 |
|  | Urban | 97 | 15 | 15.5 | 8.9 | 23.3 |  |
| School | Baleveng Evangelical Primary School | 40 | 2 | 5.0 | 0.6 | 13.5 | **<0.001** |
|  | Baleveng Central Government School | 52 | 20 | 38.5 | 25.3 | 51.9 |  |
|  | Batsinto Government Primary School | 51 | 4 | 7.8 | 2.2 | 16.5 |  |
|  | Fiala-Foreke Urban Government School | 72 | 17 | 23.6 | 14.4 | 34.0 |  |
|  | Fotetsa Government School | 61 | 4 | 6.6 | 1.8 | 13.9 |  |
|  | Foto Government School | 57 | 1 | 1.8 | 0.1 | 6.4 |  |
|  | Lepoh Government School | 49 | 3 | 6.1 | 1.3 | 14.3 |  |
|  | St Albert Catholic School Fiala-Foreke | 25 | 0 | 0.0 | 0.0 | 13.7 |  |
| Type of School | Confessional Private Schools | 65 | 2 | 3.1 | 0.4 | 8.4 | **0.012** |
|  | Government Schools | 342 | 49 | 14.3 | 10.8 | 18.2 |  |
| Age | 7-9 years | 85 | 6 | 7.1 | 2.6 | 13.4 |  |
|  | 10-14 years | 311 | 44 | 14.1 | 10.5 | 18.2 |  |
|  | 15-16 years | 11 | 1 | 9.1 | 0.2 | 30.9 | 0.204 |
| Sex | Female | 184 | 23 | 12.5 | 8.1 | 17.6 | 0.986 |
|  | Male | 223 | 28 | 12.6 | 8.5 | 17.2 |  |
| Class | Class 3 | 23 | 0 | 0.0 | 0.0 | 14.8 |  |
|  | Class 4 | 78 | 3 | 3.8 | 0.8 | 9.1 |  |
|  | Class 5 | 132 | 22 | 16.7 | 10.8 | 23.5 | **0.009** |
|  | Class 6 | 174 | 26 | 14.9 | 10.0 | 20.6 |  |
| Religion | Animist | 129 | 15 | 11.6 | 6.7 | 17.7 |  |
|  | Christian | 203 | 21 | 10.3 | 6.5 | 14.9 |  |
|  | Muslim | 14 | 2 | 14.3 | 1.8 | 36.0 | 0.150 |
|  | Other | 61 | 13 | 21.3 | 11.9 | 32.3 |  |
| Occupation of Parent | Informal sector worker | 376 | 50 | 13.3 | 10.0 | 16.9 | 0.258 |
|  | Salaried worker | 31 | 1 | 3.2 | 0.1 | 11.6 |  |
| Cohabit with domestic animals | No | 74 | 2 | 2.7 | 0.3 | 7.4 |  |
|  | Yes | 333 | 49 | 14.7 | 11.1 | 18.7 | **0.005** |
| Fowls | No | 145 | 10 | 6.9 | 3.4 | 11.5 |  |
|  | Yes | 262 | 41 | 15.6 | 11.5 | 20.3 | **0.011** |
| Dogs | No | 243 | 31 | 12.8 | 8.8 | 17.2 |  |
|  | Yes | 164 | 20 | 12.2 | 7.6 | 17.6 | 0.867 |
| Cats | No | 297 | 34 | 11.4 | 8.1 | 15.3 |  |
|  | Yes | 110 | 17 | 15.5 | 9.3 | 22.7 | 0.278 |
| Goats | No | 258 | 32 | 12.4 | 8.6 | 16.7 |  |
|  | Yes | 149 | 19 | 12.8 | 7.9 | 18.5 | 0.918 |
| Pigs | No | 278 | 25 | 9.0 | 5.9 | 12.6 |  |
|  | Yes | 129 | 26 | 20.2 | 13.6 | 27.5 | **0.002** |
| Nature of house floor | Cemented / tiled | 191 | 19 | 9.9 | 6.1 | 14.7 |  |
|  | Earth | 216 | 32 | 14.8 | 10.4 | 19.8 | 0.139 |
| Wear shoes always | No | 256 | 33 | 12.9 | 9.0 | 17.2 |  |
|  | Yes | 133 | 18 | 13.5 | 8.2 | 19.8 | 0.775 |
| Baths body daily | No | 256 | 33 | 12.9 | 9.0 | 17.3 | 0.775 |
|  | Yes | 151 | 18 | 11.9 | 7.2 | 17.5 |  |
| Keeps nails short | No | 196 | 37 | 18.9 | 13.7 | 24.6 | **<0.001** |
|  | Yes | 221 | 14 | 6.3 | 3.5 | 9.9 |  |

There was significant variation of the prevalence across health areas ranging from 1.8% (95% CI: 0.04% - 6.4%) in Fometa to 23.9% (95% CI: 15.6% - 33.1%) in Baleveng health areas p<0.001). Children attending government schools had a significantly higher prevalence of 14.3% (95% CI: 10.7% - 19.4%) compared to those attending confessional private schools with a prevalence of 3.1% (95% CI: 0.4% - 8.4%) p = 0.012. The Baleveng Central Government School topped the chart with a prevalence of 38.3% (95% CI: 25.3% - 51.9%) followed by Fiala-Foreke Urban Government School with 23.6% (95% CI: 14.4% - 34.0%), and Batsinto Government Primary School with 7.8% (95% CI: 2.2% - 16.5%) respectively. The prevalence was higher in children aged 10-14 years (14.1% (95% CI: 10.5% - 18.2%)) compared to the 7-9 years age group (7.1%(95% CI: 2.6% - 13.4%)) and those 15-16 years (9.1% (95% CI: 0.2% - 30.9%)), however, this difference was not significant, p = 0.204. There was not sex difference as the prevalence in males was 12.6% (95% CI: 8.5% - 17.2%) and that in females was 12.5% (95% CI: 8.1% - 17.6%), p = 0.986. Children in higher classes had significantly higher prevalences of tungiasis compared to children in lower classes with class 5 topping the chart with 16.7% (95% CI: 10.8% - 23.5%), followed by class 6 with 14.9% (95% CI: 10.0% - 16.9%), then class 3 with 3.8% (95% CI: 0.8% - 9.1%) and class 3 with 0.0% (95% CI: 0.0% - 14.8%), p = 0.009. The prevalence was significantly higher among children from homes with domestic animals (14.7% (95% CI: 11.1% – 18.7%)) compared to those from homes with no domestic animals (2.7% (95% CI: 0.3% - 7.4%)) p = 0.005. Although the prevalence was higher among children living in houses with earthed floors (14.8% (95% CI: 10.4% - 19.8%)) compared to those living in houses with cemented floors (9.9% (95% CI: 6.1% - 14.7%)), this difference was not significant, p = 0.139. There were no differences in the prevalence of tungiasis among children who wore shoes always or not, nor among children who took daily baths or not. However, children who kept their nails short had a significantly lower prevalence (6.3% (95% CI: 3.5% - 9.9%)) compared to those who did not tidy their nails (18.9% (95% CI: 13.7% - 24.6%)), p <0.001)

### Clinical profile of tungiasis among our participants

Fifty-one of the children surveyed were infested with the sand flea – *Tunga penetrans.* The intensity of infestation ranged from 1 to 10 fleas per infested child, with a median of 3 (IQR=2-5), for a total of 174 fleas counted in the 51 infested children. The majority (88%) of the children had mild infestations with 1-5 fleas, and 12% had moderate infestations with 6 to 10 fleas and there were no cases of severe infestation with >30 fleas (Fig 3a & b) (25). Infestation was highest in the 10-14-years age group. In the 10-14-year and the 15-16-year age groups, males were more infested than females although this difference was not significant (p=0.231) (Fig 3b). Lesions were mainly located on the toes (92%), hands (31.4%) and heels (27%) (Fig 3c). Pain (72.5%), itching (66.7%), ulceration (41.2%), skin rash (37.3%) and inflation (29.4%) were the most common symptoms, associated with tungiasis infestations (Fig 3d). Superinfections (19.6%), digital deformities (15.7%), difficulties in walking (13.7%), and abscess formation (11.8%), represented the most observed complications (Fig 3e). Among the children infested with tungiasis, 72.5% reported sleep disturbances, 49% difficulty concentrating and 23.5% irritability due to itching and pains induced by tungiasis. Some 13.7% of the children also reported at least a day of absence from school because of the infestation (Fig 3f).

**Fig 3.**
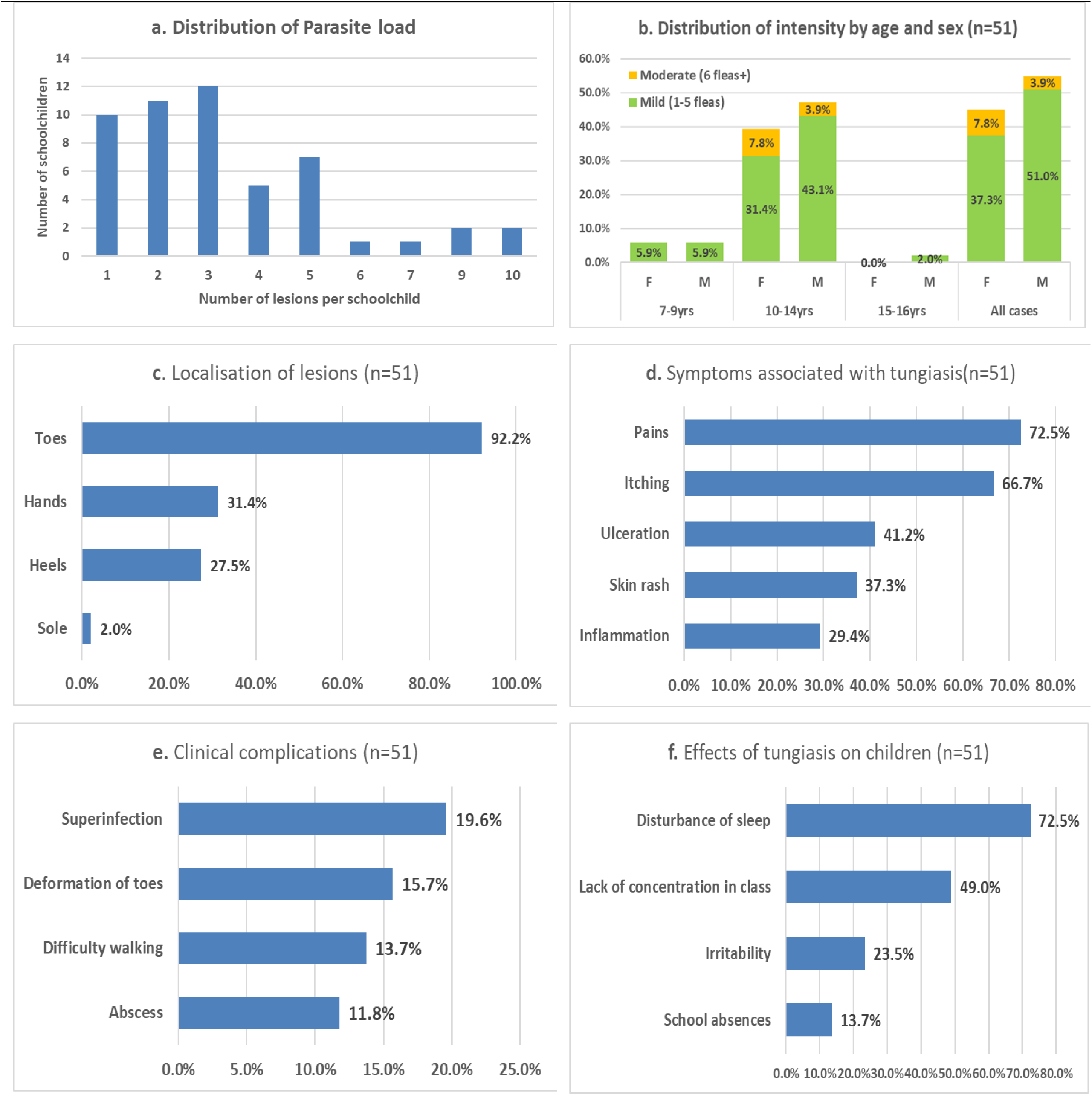
Clinical profile of tungiasis. **a.** disbution of parasite load, **b.** distribution of intensity of infestation by age and sex, **c.** localisation of lesions, **d**. symptoms associated with tungiasis, **e.** clinical complications of tungiasis, **f.** effects of tungiasis on schoolchildren.

### Treatment attitudes and practices

Fig 4 show detail of tungiasis treatment attitude and practices adopted by schoolchildren who were infested with the sand flea. Majority of schoolchildren did not consider tungiasis a health problem enough to warrant hospital consultation and treatment as only 2% declared that they would go to the hospital with their lesion, meanwhile 98% would extract their flea lesions at home. Most (84.3%) of the schoolchildren would extract the flea within 2 days of penetration. While extraction was enough for 60.8% of the schoolchildren, 21.6% would apply some ointments after extraction and 17.6% would only apply ointments to kill the flea. The major types of ointments applied were petroleum oil (vaseline), palm oil or kerosene.

**Fig 4.**
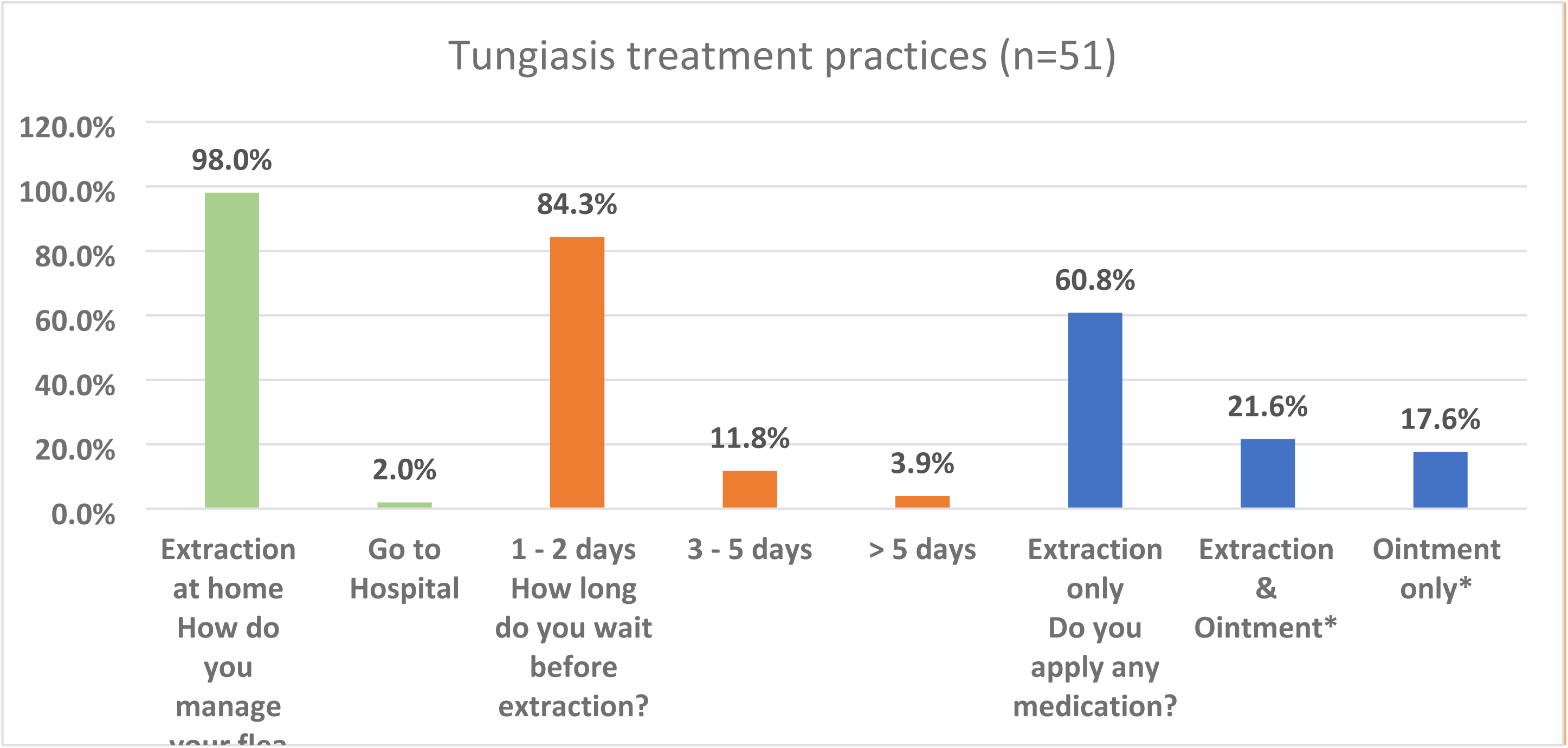
Tungiasis treatment practices adopted by infested participants. Ninety-eight percent would extract their flea lesion at home and only 2% indicated they would go to the hospital for extraction. Majority (84.3%) would extract the flea within 2 days of penetration. While extraction was enough for 60.8% of the participants, 21.6% would apply some ointments after extraction and 17.6% would only apply ointments to kill the flea. *Main types of ointments applied were Vaseline pomade, palm oil or kerosene.

### Factors independently associated with tungiasis

After the univariate analyses which revealed association of tungiasis with 8 variables (Table 2), a multivariate analysis to identify independent factors associated with tungiasis in our study, is shown in Table 3.

**Table 3.** Determinants of tungiasis among schoolchildren in Dschang Health District.

|  | OR | 95% Confidence Interval<br>(C.I.) for OR |  | P-value |
| --- | --- | --- | --- | --- |
|  |  | Lower | Upper |  |
| *Schools* |  |  |  |  |
| **Baleveng Central Government School** | **47.69** | **4.95** | **459.68** | **0.001** |
| Baleveng Evangelical Primary School | 1.50 | 0.12 | 19.14 | 0.757 |
| Batsinto Government School | 3.91 | 0.38 | 39.94 | 0.250 |
| **Fiala-Foreke Urban Government School** | **11.09** | **1.35** | **91.02** | **0.025** |
| Fotetsa Government School | 3.46 | 0.34 | 35.36 | 0.295 |
| Lepoh Government School | 2.10 | 0.19 | 22.98 | 0.542 |
| St Albert Catholic School Fialla-Foreke | 0.00 | 0.00 |  | 0.998 |
| Foto Government School | 1.00 |  |  |  |
| *Classe* |  |  |  |  |
| Classe 3 | 0.00 | 0.00 |  | 0.998 |
| **Classe 5** | **22.11** | **4.73** | **103.36** | **0.000** |
| **Classe 6** | **7.70** | **1.97** | **30.11** | **0.003** |
| Classe 4 | 1.00 |  |  |  |
| *Cohabitation with domestic animals* |  |  |  |  |
| **Yes** | **5.15** | **1.09** | **24.29** | **0.039** |
| No | 1.00 |  |  |  |
| *Cohabitation with pigs* |  |  |  |  |
| **Yes** | **2.34** | **1.13** | **4.82** | **0.022** |
| No | 1.00 |  |  |  |

Attending the Baleveng Central Government and the Fiala-Foreke urban Government Schools were independently associated with tungiasis infestation, with odd ratios of 47.69(4.95 – 459.68), p = 0.001 and 11.09(1.35 – 91.02), p = 0.025 respectively. Being a pupil of classes 5 and 6 as well were independently associated with tungiasis infestation in our study, with odd ratios of 22.11(4.73 – 103.36), p<0.001 and 7.70(1.97 – 30.11), p = 0.003 respectively. Finally, cohabitating with animals (OR = 5.15(1.09 – 24.29), p = 0.039), especially pigs (OR = 2.34(1.13 – 4.82), p = 0.022) were also found to be independently associated with tungiasis infestation.

## Discussion

### Prevalence of tungiasis and associated factors

Tungiasis, a sand flea disease caused by *Tunga penetrans* and affecting people living in shantytowns and resource-limited rural communities of Africa, Latin America and the Caribbeans, was recently considered as a neglected tropical disease by the World Health Organization(9). An informal meeting of the WHO on the development of a conceptual framework for tungiasis control held in Geneva in January 2021, recommended among other things the mapping and surveillance of tungiasis(26). We conducted this school-based study in Dschang, west Cameroon, to determine the prevalence, clinical profile and associated risks factors as part of efforts to understanding the burden of tungiasis in Cameroon. Our study is the fourth study on tungiasis so far in Cameroon following the once by Njeumi et al. (2002) in the west region(27), Collins et al. (2009) in Ndu, northwest region(11), and Bourrée et al. (2012), in Bangou, west region(23). This is indicative of the paucity of data regarding tungiasis in Cameroon, in line with the situation in the whole of sub-Saharan Africa echoed by Obebe et al.(2020) (1).

Our study revealed an overall prevalence of 12.5% (95% CI: 9.5% - 15.9%) among the schoolchildren 7-16 years of age in Dschang health district (Table 2). The prevalence in our sample was lower compared to the 60.5%-72.1% and 31 – 40.9% found in similar age-groups in Ndu, northwest (11) and Bangou, west(23) Cameroon respectively. It was equally lower than the pooled prevalence of 31.89%, from 23 studies carried out among school children in 8 countries of sub-Saharan Africa(2). Although seasonal variation in prevalence has been reported with lower prevalence rates noted in the rainy season compared to the dry seasons(12,17,28), the relatively lower prevalence in our study couldn’t be attributed to seasonal variation given that our study was conducted in April and May at the beginning of rainy season, similar to the studies in Ndu and Bangou that were respectively conducted in august(11,23), in the heart of the rainy season. Equally, Dschang lies in the northwestern highlands of Cameroon as Ndu and Bangou that all share the same climatic conditions and therefore, could not have influenced the variation in prevalence of tungiasis among these localities. The 10-14-year age group had a higher prevalence (14.1%(95% CI: 10.5% - 18.2%)) compared to the 7-9-years (7.1%) and 15-16-years (9.1%) though this difference was not significant. This corroborates findings in Kilifi-Kenya(29) but differs from findings in Ndu-Cameroon and Wensho-Ethiopia where prevalence was higher in the 5-9year age group(11,13). There was no gender difference in the prevalence of tungiasis in our study unlike in many other studies including Ndu-Cameroon(11), Wensho-Ethiopia(13), Oromia-Ethiopia(30), Kilifi-Kenya(29), and in Fortaleza-Brazil(12), where there was a male predominance, in prevalence and infestation intensity. The prevalence was significantly higher among children in classes 5 and 6 compared to children in lower classes (p = 0.009). Children attending government schools had significantly higher prevalence compared to those attending private confessional schools (p=0.012). Generally, in Cameroon, private confessional schools have better infrastructure where classrooms have cemented floors and walls, and children are inspected regularly for cleanliness. This contrasts with most government schools, especially in the rural areas where school infrastructure is usually uncompleted or has poor maintenance, and where there is a laisser-faire attitude regarding personal hygiene. The classroom floors of the Baleveng Central government schools and Fiala-Foreke urban government schools where the prevalence of tungiasis were highest in this study, were cracked, while the classroom floors of St Albert Catholic school where there were no schoolchildren with tungiasis had smooth concrete floors (Fig 2). This corroborates findings in Kilifi-Kenya where natural soil/sandy or cracked concrete classroom floors was associated with higher prevalence of tungiasis among schoolchildren(15). Children from homes where there was cohabitation with domestic animals had a significantly higher prevalence of tungiasis (p=0.005), and this was more prominent if there were fowls (p=0.011) or pigs (p=0.002). This confirms findings from studies in sub-Sharan Africa(1,2) and South America(5,10,31).

Three factors were found to be independently associated with tungiasis in our study (Table 3). Cohabitation with domestic animals increased the odds of tungiasis infestation by 5.15 folds, meanwhile children cohabiting with pigs particularly were 2.34 times more likely to be infested. This finding was in line with that in Bangou, west Cameroon(23) and in other sub-Saharan countries(1,2,13,29), and confirms tungiasis as a zoonotic disease where domestic animals constitute reservoirs(3,7,32). In our study, children attending government schools were more at risk, especially those attending the Baleveng central government school and the Fiala-Foreke urban government school, where school children were 47.69 and 11.09 times respectively, more likely to be infested with tungiasis (Table 3). The classroom floors of these two schools were cracked concrete, and confirms the findings in Kilifi-Kenya, where the odds of tungiasis infestation were higher in schools where classrooms had natural sand/soil or cracked concrete floors(29). The risk factors identified in this study provide some evidence that could be explored for the development of a control strategy in Cameroon, learning from the experience of Kenya(33).

The parasite load in schoolchildren infested with *Tunga penetrans* in our study was relatively low, ranging from 1 – 10 fleas per infested child. Majority (88%) of the children were mildly infested with 1-5 fleas and 12% moderately infested. Our findings are comparable to the findings in Bangou, west Camerron, where 84.7% and 13.9% of participants had mild and moderate infestations with tungiasis respectively(23), but differs from those in Ndu northwest Cameroon(11), kilifi-Kenya(29) Wensho-Ethiopia(13) and Igbogoda-Nigeria(14) where the intensity of infestation were higher. The pattern of infestation intensity in our study was similar to those described other studies in sub-Saharan Africa and South America where the greater majority were mildly infested and a few with heavy infestations(5,11,14,25,29). This pattern could provide a control option in resource-limited situations by concentrating efforts on the few individuals with heavy infestations, who are at the same time the perpetrators of tungiasis transmission(25).

### Tungiasis morbidity and treatment practices

The rate of infestation was highest in the 10-14-year age group in both male and female, though with a slight male predominance (Fig 3b). This is inline with most studies in sub-Saharan Africa(2). This has been explained by the fact that children in this age group play a lot both in school and at home, and most often do so barefooted on dusty fields, thus exposing themselves more to the fleas(14,21,34). The difference in infestation between boys and girls have been attributed by some researchers to poorer observance of personal hygiene by boys compared to girls of the same age groups(21). However, others hold that boys play and move about more that girls and therefore expose themselves more(25).

As expected, most of the lesions were located on the toes. Toes have been described by most studies as the area of predilection for *Tunga penetrans*(5,12,18,25). The heels and the sole of the feet were other locations in infestation in our sample (Fig 3c). Pooled together, most lesions occurred on the feet. By nature, *Tunga penetrans* do not jump very high, and consequently, lesions are mostly limited to the feet(5). However, in our study a reasonable proportion of lesions were also located on the hands. This is not uncommon as locations of lesions other than the feet have been shown in other studies(14,24). The occurrence of lesions mostly on the feet indicates that control strategies focused on feet hygiene and feet protection can greatly reduce the prevalence and the intensity of infestation with tungiasis.

The major clinical manifestations of tungiasis infestation in our study were pains, itching, ulceration, skin rash and inflammation/swelling (Fig 3d). These same manifestations have also been reported in other studies(14,24). Itching, a primary symptom, led to constant scratching of the lesion site, provoking scratch marks or desquamation around the site. The manipulation of the lesions with sharp objects (sharpened bamboo sticks, needle, pins…) during extraction of the flea led to ulceration and inflammation, exacerbating pains at the lesion sites. The schoolchildren reported carrying out the extraction by themselves or by the help or the parents or siblings. Most of the time, no aseptic measures like cleaning the lesion with water and soap and/or topical antiseptics were applied before the extraction of the flea, thereby exposing the lesion to superinfection and abscess formation, seen in 19.6% and 11.8% respectively of the infested children in our study (Fig 3e). Superinfection is a common complication in tungiasis infestation(5,14,20,24). Chronic infestations with tungiasis led to other complications like deformation of toes and difficulties in walking among some of our participants (Fig 3e). This confirms findings from studies conducted in sub-Saharan Africa and South America(14,21,24). Quality of life was negatively impaired by tungiasis infestation among our participants. The 72.5% sleep disturbance among our participants (Fig 3f), was above the 19.2% reported in southern Nigeria(14) but close to the 86% reported among children in Kilifi-Kenya(21). On the other hand, the difficulties in concentration found in 49% of our participants was substantial although less than the 84% reported among children in Kilifi-Kenya(21). Most of our participants attributed sleep disturbance and lack of concentration to itching and pains caused by tungiasis infestation, corroborating findings in Kenya(21). School absenteeism was declared by 13.7% of our participants. In a Kenyan study, tungiasis infested children had 1.49 more odds of absenting from school compared to uninfested children(22), and consequently poorer school performances among infested children.

Most of our participants (98%) did not consider tungiasis as an important health problem that should warrant visiting a health facility for treatment and would therefore carry out extraction of lesion at home (Fig 4). This attitude and practice that potentially exposes the infested children to superinfection and deadly complications like tetanus(6) are contrary to current WHO treatment recommendations(35). Furthermore, in addition to extraction some of infested participants in our study applied some ointments like vaseline oil, palm oil, and kerosene. These were less likely to favour rapid healing but could increase inflammation leading to increased pains and higher risk of superinfection especially for kerosene applications. The WHO recommends application of low-viscosity dimeticon for the treatment severe tungiasis infestation and combination of surgical extraction and application of low-viscosity dimeticon for mild infestation, and coconut oil or neem oil for all forms of tungiasis infestation in the absence of dimeticon(35).

### Limitations of the study

The schools selected for the study were visited by for data collection only once and only schoolchildren present on the day of the passage of the survey team were recruited. Furthermore, schoolchildren below class 3 were not included in the study. These might have underestimated the prevalence of tungiasis in our study.

## Conclusion

Our study reveals that tungiasis is substantially prevalent, affecting 12.5% of primary schoolchildren in the Dschang health district of Cameroon. The presence of tungiasis was associated with Cohabitation with domestic animals especially pigs, attending government schools and being a pupil of classes five and six respectively. Tungiasis orchestrated substantial morbidity and affected the quality of life in infested schoolchildren. The schoolchildren adopted poor tungiasis treatment attitudes and practices contrary to the current WHO treatment recommendations. We therefore recommend education of household heads on the need for confining domestic animals especially pigs into separate spaces (piggery) and regularly spraying these animals and their confines with insecticides; as well as insisting on personal hygiene for their children; adopting the WHO recommendations of limiting home extraction of tungiasis lesion and applying of low viscosity dimeticon or neem/coconut oil to lesions instead. Equally, education of the schoolchildren on the need for personal hygiene including regular wearing of shoes, and schoolteachers on regular hygiene inspection of schoolchildren, and watering of classroom floors. More studies are required to understand the wider spectrum of tungiasis disease in Cameroon.

## Data Availability

All the data is provided in the manuscript and attached files

